# Protocol for the pilot study of group video yogic breathing app in breast cancer survivors

**DOI:** 10.1101/2022.04.08.22273610

**Authors:** Sundaravadivel Balasubramanian, Jennifer Harper, Katherine R. Sterba, Ramakrishnan Viswanathan, Harriet Eldredge-Hindy

## Abstract

**Introduction:** Breast cancer remains a leading cause of cancer deaths; however, recent improvements in treatment have improved survivorship. As a result of this improvement, more individuals are living with the long-term side effects of cancer treatment. Therefore, methods that incorporate lifestyle and mind-body approaches are becoming increasingly used in the patient treatment pathway.

**Methods:** In this study, PranaScience Institute will develop and test a group video mobile application for Yogic Breathing (YB). YB is shown to reduce symptomatic conditions associated with several conditions including breast cancer. For this initial feasibility study, PranaScience will collaborate with the Medical University of South Carolina to implement the study app-based program in breast cancer survivors. This research is aimed to understand if the YB could be delivered via an app, if participants are able to practice it satisfactorily, and if there is any symptom relief by the YB practice. In the control group, participants will be directed to the Attention Control (AC) feature of the app, which guides users to focus on a mindfulness activity not involving YB. Participants will be randomly assigned to the YB or AC study plan (N=20 per group). Breast cancer survivors who have completed radiation therapy within last 6 months will be recruited for this study and provided access to the app for a 12-week program. The study app will record total practice times. Virtual visits by a study yoga instructor during group video sessions will measure participant compliance with proper technique. Feasibility will be examined by evaluating intervention delivery factors and resource needs. Acceptability of using the mobile study app to support symptom management will be evaluated using a satisfaction and system usability scale. Behavioral survey measures will help guide effect sizes and power calculations for the next larger-scale study. Biomarkers in the saliva (tumor suppressors, cytokines), and fingernails (cortisol, differential proteomics) will be measured at baseline and end of study at 12 weeks.

**Discussion:** All findings from this pilot study will be synthesized to refine the mobile study app in preparation for large-scale evaluation in Phase II involving all-study site participants with cancer. **ClinicalTrials.gov Identifier NCT05161260**.

## INTRODUCTION

### Cancer survivorship is associated with long-term chronic medical issues

Cancer is the second leading cause of death in the US, with breast cancer being the most predominant type for cancer in women[1]. Due to improvements in early diagnosis and treatment, the number of cancer survivors in the U.S. has steadily increased[1]. Approximately 90 percent of those diagnosed with breast cancer now survive five years after diagnosis. For instance, in 2019, there are more than 3.8 million breast cancer survivors in the US[2]. As cancer becomes a chronic condition rather than a life-threatening illness, the health care community must learn to recognize and manage the long-term sequelae of physical and psychological side effects of treatment protocols. Major issues receiving the most attention include chronic inflammation[3, 4], increased risk of second primary malignancies[5], pain and muscle weakness[6], depression and anxiety[7–10], fatigue[11], sleep disturbance[12], dryness of the mouth[13, 14], and less threatening medical conditions. In addition, preexisting comorbidities such as obesity, hypertension, hyperlipidemia, and diabetes will most certainly affect disease-free survival, and ultimately, overall survival[15]. For example, breast cancer survivors are at significantly higher risk for developing anxiety and depression compared to women who do not have breast cancer[16–19]. Managing these symptoms is essential, as mortality rates were 39 times higher in patients who had been diagnosed with major depression[7], and a decrease in depression symptoms was associated with longer survival in metastatic breast cancer patients[20]. As a result of this changing landscape, survivorship care programs provide a critical component of the patient treatment pathway. ***Looking at improving and sustaining long-term survival in cancer requires a total-health strategy that includes a focus on non-pharmacological lifestyle and mind/body interventions***.

### YB alters salivary proteome

The molecular mechanisms of YB in isolation have not been studied in detail[54], and there are currently no established protein biomarkers that can be used to evaluate the effects of YB on clinical outcomes or wellbeing. Saliva is an easily accessible source of biological material that can be used to assay biomarkers in a non-invasive manner[55–58]. Further, salivation is one of the parasympathetic activation responses[59]; thus, we hypothesized that a proteomic survey of saliva following YB may reveal proteins that are altered in response to YB. Twenty healthy volunteers were randomized to a YB or Attention Control (AC) group in collaboration with a biostatistician to ensure equal gender distribution. YB participants were tested one-on-one with a yoga instructor who taught participants how to perform YB, which included two phases, namely, Chanting Om and Thirumoolar Pranayamam (TMP), each for 10 min. AC participants were guided to perform quiet reading for 20 min in a seated position in presence of yoga instructor. Saliva samples were collected from participants in both groups at 0, 5, 10, 15, and 20 min. Protein was extracted and quantified from saliva samples and 100 μg analyzed by LC-MS/MS to identify differences in protein composition driven by YB. Comparison of proteins following YB or AC using a normalized spectral counting approach revealed significant increases in a number of proteins involved in immune response and tumor suppression including deleted in malignant brain tumor 1 (DMBT1), which has been shown to be downregulated in many cancers[60], and IGLC2, which is known to be increased during viral infection and thought to play a role in immune response[61, 62]. Targeted western blots were used to validate these findings [39]. ***These findings represent the first demonstration that YB induces changes in biomarkers of immune response and cancer***.

### YB reduces levels of pro-inflammatory proteins

Yoga practices are widely perceived to be stress-reducing; however, the molecular mechanisms for how yoga practices such as YB affect stress response are lacking. In a study of twenty healthy volunteers stratified by gender into a YB or AC group (n=10 per group), we examined the effects of YB on levels of salivary cytokines. As in the aforementioned study, enrolled participants performed a 20 min YB or AC intervention one-on-one with a trained instructor and salivary samples were collected at 5 min intervals from both groups. Cytokine multiplex assays were performed on the samples to look at ten stress-related cytokines (IL-1β, IL-1A, IL-6, IL-8, IL-10, IL-17, IP-10, MCP-1, MIP-1b, and TNF-alpha). Levels of the cytokines IL-1β, IL-8, and MCP-1 were significantly lower in the YB group. IL-1β is implicated in stress, inflammation and pain perception[63, 64], and is currently being pursued as a clinical target for autoinflammatory syndromes[65]. IL-8 is a key mediator of inflammatory processes during various disease conditions involving oxidative stress; elevated levels of IL-8 are associated with poor prognosis in breast cancer[66]. There are currently no therapies that directly target IL-8. MCP-1 is elevated and secreted in response to chronic inflammation and is a biomarker for coronary heart disease[67]. ***These early studies, though correlative, provide the first evidence that breath regulation, even for short periods of time, have potential to induce measurable changes in biological functions related to stress response and inflammation***.

### YB in-person group practice is an acceptable and feasible intervention for cancer patients and Caregivers

To evaluate whether cancer patients and their caregivers would be receptive to a YB program for symptom relief and support, the PI conducted a pilot study at the American Cancer Society Hope Lodge in Charleston, SC over a 9-month period. The Hope Lodge facility provides temporary housing to cancer patients and caregivers that are receiving treatment at a nearby cancer treatment facility. Guests were recruited to participate in a YB class consisting of 20 minutes of YB exercises. For each class, a certified yoga instructor provided demonstration and verbal instruction for five different YB exercises to a group of 2-5 participants. At completion of the class, participants were surveyed via a voluntary questionnaire to assess sociodemographic (age, gender) and clinical (cancer type) characteristics, perceived and experienced YB benefits (mood, pain, appetite, among others), and likelihood of the patient practicing these methods in the future. Responses to open-ended questions were reviewed using a group process to identify themes. Patients of all cancer types attended classes at rates comparable to control caregivers with head and neck cancer patients participating most frequently (27.8%), followed by breast (13%) and brain cancer (9.3%). No adverse effects were reported by any of the patients. All participants were very (90%) or somewhat (10%) satisfied with the experience, with the majority comfortable with the session length (83% reported length was “just right”; 17% reported “too short”). Most participants (83%) reported that they would continue to do exercises at home. Majority of participants indicated that they would be very (46%) or somewhat (40%) interested in participating in a longer program if offered at a convenient location. ***Self-reported measures collected from participants indicated perceived improvements in stress, pain, and mood after a single YB group session***. Open-ended comments from participants highlighted some general themes focused on impact, logistics, resources/ marketing. Additional data specifically from women with breast cancer (Survivors’ Fit Club, a 10 wk group program) reflected similar self-reported and thematic outcomes (Support letters from two breast cancer survivors who attended the YB sessions are attached). Overall, cancer survivors rated YB experiences favorably and expressed enthusiasm for continued practice if given the appropriate resources. Finally, PranaScience has been a leader in providing group video YB sessions via Zoom app for various target audiences since the year 2018.

## METHODS

### Intervention to be studied

| Intervention Type | Name | Description |
| --- | --- | --- |
| Behavioral (e.g., Psychotherapy, Lifestyle Counseling); | Pranayama-based group video app for yogic breathing | The YB app (alpha version) will be a first production version of a Yogic Breathing mobile app that guides users through proscribed YB exercises version and contains special features designed to measure compliance and capture feedback from the user in the Phase I program. |
| Behavioral (e.g., Psychotherapy, Lifestyle Counseling); | Attention Control app feature | An Attention Control (AC) mode will be integrated into the app so that users can operate the app in AC mode instead of YB. "AC mode" will be visually and functionally similar to the YB mode to avoid variability due to different user experiences. In place of the YB session, there will |
|  |  | be a similar session providing instruction on an AC activity, which will be presented to the user as a "mindfulness practice". This will serve as a control for YB and will not be carried into the commercial version(s) of the app. |

#### Group video practice app for peer-supported home-based practice of Yogic breathing

Complementary and integrative health approaches are increasingly being used by cancer patients and survivors to manage symptoms that do not always respond to pharmacological interventions, such as stress, mood, and fatigue[21]. In this regard, yoga therapy, which encompasses a constellation of mind-body practices including meditation, physical exercises or postures, and breath control, has emerged as a valuable complementary therapy for cancer patients and survivors[22–29]. Yogic breathing (YB or *pranayama*) consists of a collection of techniques used to voluntarily regulate breathing without the need of physical exertion[30]. YB induces a strong relaxation response via vagal and other parasympathetic stimulation[31, 32] which has been causally linked to genes involved in the inflammation, stress response, metabolism, and tumor suppression[33–36]. We demonstrated that YB stimulates <u>*1)*</u> reductions in levels of salivary pro-inflammatory biomarkers[37], <u>*2)*</u> induction of the neuromodulator Nerve Growth Factor (NGF), which is important for maintenance and survival of the peripheral and central nervous system[38], and <u>*3)*</u> alterations in the levels of proteins associated with tumor suppression and immune regulation[39]. Although mobile Apps provide a convenient way to provide mindfulness exercises and could reduce barriers of delivery intervention[40–42], adherence to such apps is at very low rates [43, 44]. However, sustained practice of yoga [45] alongside other behavioral interventions (diet, exercise) are considered feasible, acceptable, and successful through peer support where the participants motivate each other for enhanced adherence, and improved outcomes[46, 47]. Peer support is recognized an effective framework for sustained adherence to interventions in cancer[48–51]. In this context, we envisaged that group video chat apps[52, 53] could provide a unique opportunity to deliver YB interventions for simultaneous practice by participants in virtual groups. While additional work is needed to demonstrate the efficacy of specific mind-body approaches in improving health outcomes for cancer survivors, *there is clear opportunity for the development of new tools that promote and facilitate acceptable, feasible, and sustainable mind-body intervention strategies* that serve to improve overall wellbeing. The goal of this Small Business Technology Transfer Research (STTR) project funded to PranaScience by the National Institutes of Health (NIH) is to develop an *app for group video practice of YB that is optimized for home-based delivery of YB programs* and maximally effective in reducing symptomatic conditions associated with cancer treatment and survival. For the Phase I portion of this project, PranaScience will build a prototype mobile app and test the app for adherence, feasibility, and acceptability to support symptom management in a pilot study of breast cancer survivors in collaboration with the Medical University of South Carolina (MUSC) and a mobile technology firm.

### Study Endpoints

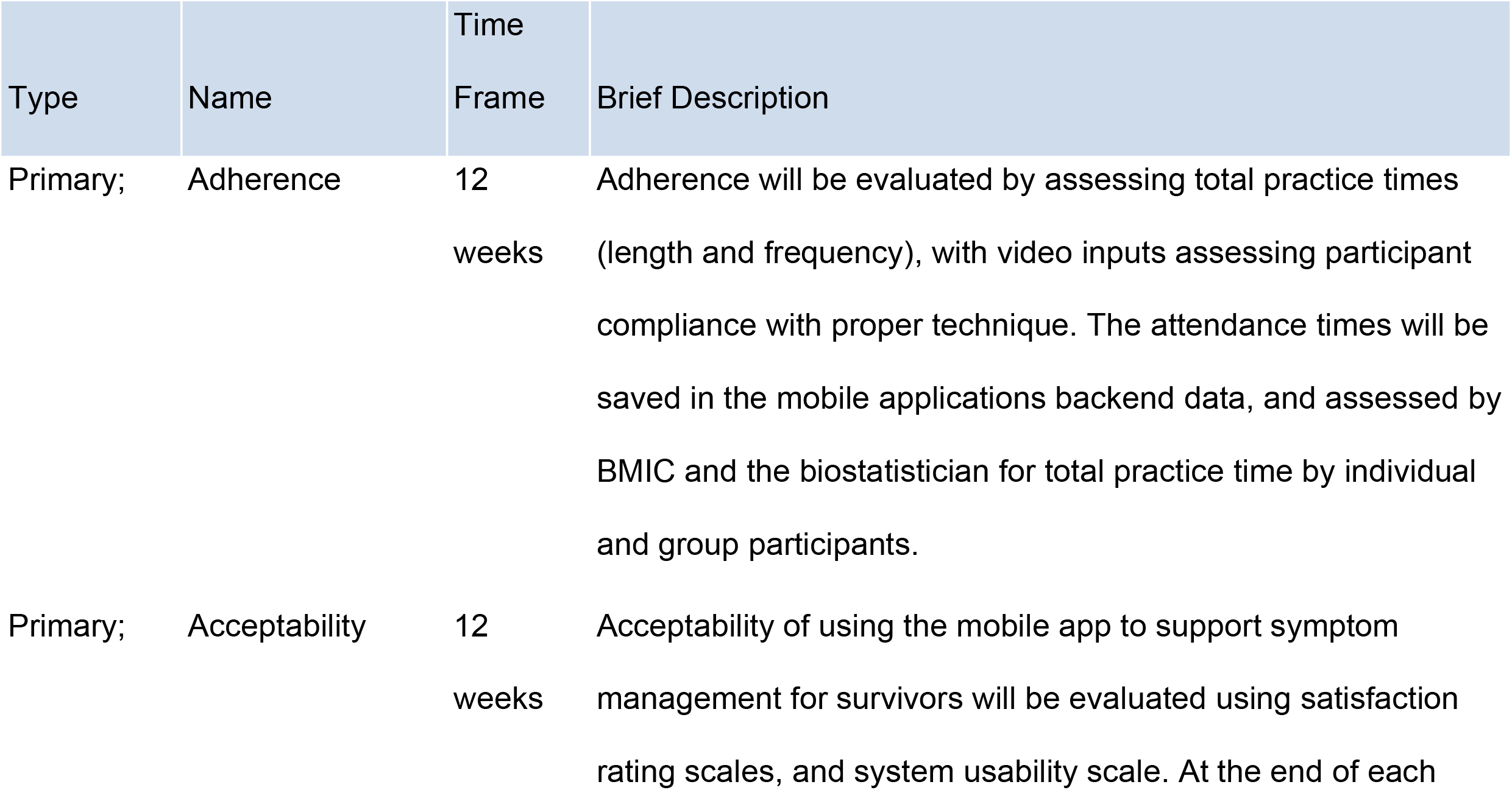
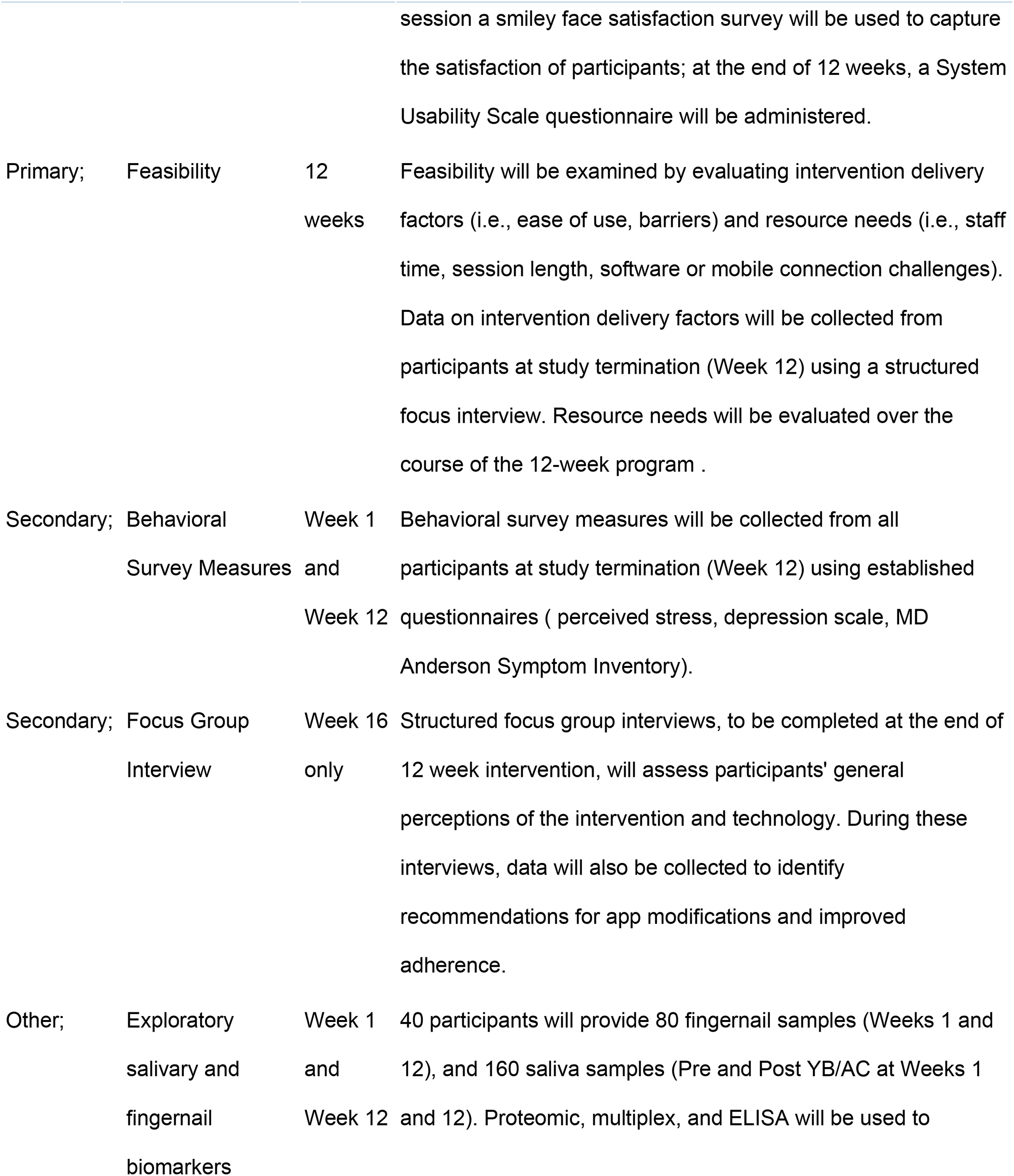
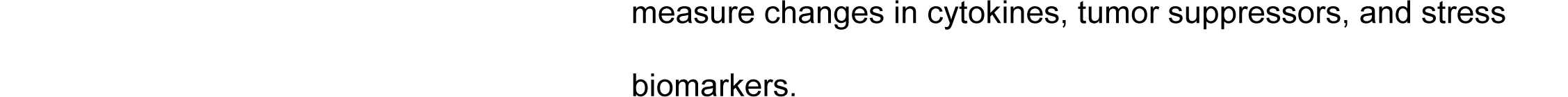

### Inclusion and Exclusion Criteria/ Study Population

All Radiation Oncology staff at MUSC are cross trained in research referral to study staff for all potential participants, and research staff are closely connected to the clinic staff. Potential participants will be screened by the study radiation oncologist (PI) and study coordinator using oral questions and medical records about their diagnosis, radiation treatment and willingness to participate in the study.

### Inclusion criteria

1. Age 18 or older, willing to give informed consent
2. Diagnosis of Stage 0-III breast cancer
3. Completion of radiation therapy within last 6 months
4. ECOG performance status of 0-3 during cancer treatment
5. Orally expressed visual and auditory acuity adequate for filling out all study forms and participating in telephone interviews and internet-based group video applications
6. Access to telephone, and internet-connected computer or mobile phone.

### Exclusion criteria

1. Subject is unwilling or unable to comply with any of the study procedures
2. Orally expressed current dependence on alcohol or drugs

### Inclusion of Women and Minorities

This study will include only women, as current breast cancer survivors are predominantly women. The inclusion of minorities in breast cancer survivorship is recognized as being of critical importance due to increased mortality and morbidity among African Americans. Based on our preliminary studies from Hope Lodge, and Survivors’ Fit Club, we will have minority representation as follows: 35-50% African American, 8-10% Hispanic, and 4-6% Asian American. Based on our past success in recruiting individuals from diverse racial and ethnic groups, we do not anticipate problems with hitting our target enrollment numbers from these groups and at rates proportionate to their representation in our catchment area. Existing Datasets or Resources. None to report.

### Inclusion of Children and Individuals Across the Lifespan

Women 18 years and above will be included in the study. There is a very low occurrence of breast cancer among women under 18, and thus no children will be recruited in the present study.

### Number of Subjects

A total of 40 breast cancer patients will be recruited for this study. This pilot study will enroll a total of 40 study participants, 20 subjects per study arm. Given this sample size, we will be able to estimate differences between the groups within a margin of error of 14.2% with 95% confidence level.

### Setting

The study will be centrally managed by PranaScience Institute, headed by Dr. Sundar Balasubramanian, and the subjects will be referred to the study by the sub-award PI Dr. Harper Professor, Dept. of Radiation Oncology at the Medical University of South Carolina (MUSC). PranaScience offices, located near the Medical University of South Carolina campus in downtown Charleston, SC. Mindfulness instructor Katharine Hendrix works with the PI on a number of research projects. For this study, she will act as a consultant to provide yogic breathing instruction to study participants at study initiation, and weekly support thereafter.

Consent processes, yogic breathing instruction, app demos, collection of patient surveys and focus group interviews, as well as all data meetings, will take place at PranaScience office facilities in dedicated office and conference spaces, or completed remotely via telephone, video conferencing applications and electronic data collection instruments. The physician investigators will advise the study team on breast cancer diagnoses, radiation treatment, side effects, disease prognosis, ECOG performance status, and management of symptoms. Physician investigators will also help with recruiting and informing potential study participants about the research study. Dr. Sterba, Associate Professor, Department of Public Health Sciences, MUSC, will conduct focus group interviews along with the study coordinator at the PranaScience conference room and collate the data from the interviews. The PI will have biweekly meetings with the research team including the research coordinators at SCTR, Dr. Balasubramanian and Dr. Hendrix through email or phone calls.

### Recruitment Methods

#### Recruitment

Charleston, SC, home of the PranaScience Institute and Medical University of South Carolina (MUSC), Hollings Cancer Center, along with the Roper Breast Care Center, is the site of a large number of cancer treatment outcome studies each year, including several studies on breast cancer. As such, all Radiation Oncology staff are cross trained in research referral to study staff for all potential participants, and research staff are closely connected to the clinic staff. Specifically, this study will use the following recruitment plan, which involves 6 potential entry paths to the project.

Following IRB approval, 1) an information brochure advertising the study will be mailed to the growing MUSC roster of over 1,500 breast cancer survivors; 2) recruitment flyers will be placed in prominent locations in MUSC, the Roper Breast Care Center, Hope Lodge, MUSC Wellness Center and clinics within the Charleston metro catchment area; 3) the study will be posted on the MUSC “active research projects” web page, at SCresearch.org, and on ClinicalTrials.gov; 4) the study will be advertised via weekly newspaper research recruitment sections (MUSC) and on social media, particularly in facebook groups of cancer survivors and support groups; 5) patients may be referred to the study by the participating investigators/clinicians; 6) a series of advertisements will be run in relevant local newspapers if the first 5 strategies are ineffective. The PranaScience Institute, under Dr.

Balasubramanian’s direction, provides active breathing exercise classes to several target patient populations including breast cancer patients and survivors.. Potential participants will be screened by the study coordinator and the PI, and the recruitment letter that contains the contact number of the study coordinator will be sent to the potential participants. Those that are interested will call the number for confirming their eligibility. The eligibility will be confirmed by oral answers provided by the participants over the telephone or their medical records reviewed by the PI and the coordinator. The coordinator will then schedule an appointment for the first visit (week 1) at the PranaScience office. At the first visit, the informed consent procedure will take place. The participants will then be randomized to condition by the study coordinator using the randomization protocol designed by the study statistician (Dr. Ramakrishnan).

#### Consent Process

Eligible participants that are screened by the study coordinator and the PI will be contacted by the recruitment letter sent by the PI. Potential participants can contact the study coordinator directly from the details presented in the recruitment advertisements. Those that are eligible will be scheduled by the Study Coordinator for the first visit at which the written informed consent will be obtained from the participants. This visit will take place at the PranaScience facility at a time mutually agreed upon by the participant and the Study Coordinator, or completed remotely via telephone with the use of an electronic consent. In a private one-on-one setting the Study Coordinator will explain the study details including the goals of the study, number of participants, number of visits by each participant, required exercises to be done on the video chat using the mobile application, biological samples to be collected, focus group interviews, risks and benefits to the participant, and monetary compensation for participation. The participant can take their time to read the Informed Consent Form in entirety and ask questions to clarify any questions that they may have. Once the participant is satisfied with the details of the study information they will sign the Informed Consent Form. A copy of the signed Informed Consent Form will be given to the participant by the Study Coordinator. After consent is obtained, participants will be randomized to YB or AC group and they will be progressing to the week 1 study visit procedures on a one-on-one basis with the study coordinator and the instructor. There is no waiting period between signing the consent and the baseline measurements.

#### Study Design / Methods

After consent is obtained, participants will be randomized by the study coordinator to YB or AC group and the study coordinator will provide the link to download the YB or AC app depending upon the group assignment. Any technical assistance in downloading the app will be provided by the study coordinator. For privacy and health reasons, the study coordinator will not touch the participant’s mobile device at any time. The coordinator will have a separate device for demonstrating the download procedures. At study initiation (Week one) at PranaScience location, participants will complete the baseline behavior assessment measures, medical history, demographics review, and will provide fingernail clippings.

Baseline scores will be established for all measures. Participants who are unable or unwilling to complete the baseline visit in person may be able to complete the baseline visit remotely. If the visit is conducted remotely, study staff will use telephone or video conference applications to meet with the participant and provide instructions for the visit. Participant surveys will be sent to the participant as electronic surveys to complete. A sample collection kit will be sent to the participant prior to the visit with instructions on how to collect and ship samples. Participants will also take part in a one-on-one training session with the mindfulness instructor Dr. Katharine Hendrix to familiarize them with the app features and the proscribed AC or YB exercise. Passive drool whole saliva will be collected (2-5 mL) before and after the AC or YB exercise. For the remote sample collection, the saliva will be collected using FDA-approved filter paper discs shipped to the participants. Instructions will be provided to collect and ship the saliva back to the PI’s lab.

The mindfulness instructor will demonstrate how the App will work to facilitate the group sessions. For example, when a YB group participant opens the App and enters into the virtual meeting, the App will notify all the other participants from within the YB group that a participant from their group is available to practice together. This step will motivate other participants to join the meeting room. The participant can choose to practice alone or choose to allow more participants (2-7 participants allowed). The participants can also ‘lock’ the meeting space when all of them within the room decide to lock the room so they can start the practice without interruption. They can choose to keep it open so others can join. The App will have a timer to start the 10 minutes practice and will ring a bell at the end of 10 minutes. At the end of the practice, a smiley face satisfaction survey will appear for individual participants that they will be asked to submit by single click. The participants can stay in the room to chat or leave or continue with longer practice. The App will count the practice times as those in between the timer bells. The App will also capture information on how long and how frequent a participant accesses the App. The similar procedure will happen for the AC participants where the YB exercise will be replaced with a mindfulness practice. While on study, each participant will be engaging in group video AC or YB practice upon initiation based on the notification by the app or a group member. There will be 3-4 subgroups for each AC or YB group each containing 5-7 participants. If a participant does not access the app for more than 24 hours during weekdays (Monday through Friday), then the App will send a reminder notification to motivate them to practice (For example, *“Hello XXX, We hope you are doing well. We have not seen you practicing the mindfulness exercise yesterday. We understand you may be busy. We hope you can catch up with the practice when you get a chance. Have a good day!”*). The motivational messages, reminders and any other text messages sent from the App to the participants will be appropriately tailored by the study biobehavioral co-investigator Dr. Sterba.

Each YB session will open with a short question-answer to assess (a) what motivated the user to open app, (b) when was the previous large meal, and (c) which nostrils are open/blocked. Participants that answer that both nostrils are blocked will be recommended to try the exercise at a later time when they are able to breathe freely. Once the participant enters into the group meeting area everyone can see and hear the other participants. To protect privacy, the participants can use any privacy protection methods such as renaming, wearing eye masks, replacing their images with other images (for example, *Avatars* on zoom), and turning their video off. However, when the mindfulness instructor is present to assess the exercise compliance by participants on a random basis, then the video has to be turned on to make sure that the participant uses the right technique. These options will be explained to the participants during the introduction of the App by the study coordinator. The provider window will show YB instructional video or visual/audio cues chosen by the group. Additional support videos will be provided via an online link that can be accessed for reference by the users. All those in the group will practice the YB exercise for 10 minutes. At end of session, a second Question and Answer (Q&A) session will assess (a) satisfaction using a facial expression scale, and (b) offer the option to set a reminder for the next practice.

AC sessions will be identical in format to YB sessions except for following differences: (a) recorded session with instructor will guide the user though a mindful exercise. Namely, user group will be guided to close their eyes or to focus on the screen (same visual displays and music will be used for YB and AC modes). No instruction will be given related to breathing or breath control. While on study (Weeks 2-11), Dr. Hendrix will join the virtual group exercise sessions on a weekly basis to each group to encourage compliance with study protocol and collect data regarding barriers. The app will additionally employ motivational nudges and reminders throughout the study.

At end of study visit on week 12, participants will complete post-study behavioral assessment measures and a system usability scale. Fingernail and saliva samples will be collected similar to baseline. As with the Baseline visit, this visit may be completed in-person at the PranaScience location, or remotely. After completing the study, participants will be asked to participate in a 60-minute, audio-taped focus group interview at (N= 2 groups; one for each study arm [AC or YB]) that will assess their perceptions of the program and technology (i.e., ease of use, recommendations for app modifications and improved adherence). The focus group will be completed via video conference approximately 1-6 months after a participant completes the study.

From the mobile App, data such as how many times the participant opened the mobile App, how long was each practice, and if the practice involved group or individual practice will be collected. No study data will be stored in participants’ devices. All the data will be transmitted to secured cloud storage and the Medical University of South Carolina’s Biomedical Information Center (BMIC) where it is stored securely. The participants will be able to access their personal data on the history of their practice.

Subjects will be required to agree to Terms of Use or EULA or Privacy Policy document (to be provided to the IRB in the future) at the time of downloading the App. During the first clinical visit the research coordinator will provide the participants a link from where the participants will download the App. The App will work on iPhones, Androids, and on desktop/laptop computers. The app is not available on app stores. Once the study is completed, at the visit 2 (week 12) the participants will be asked to delete the App. They will no longer be able to access their data from BMIC.

#### Specimen Collection and Banking (if applicable)

Saliva samples and fingernail samples will be collected at the PranaScience location by the study coordinator from participants for protein and stress hormone analysis.

#### Saliva

Saliva (about half a teaspoon) will be collected by passive drooling into a collection tube or by using filter paper discs at four time points as described below:

Saliva Sample 1: Visit 1 on week 1. Before the study exercise. 0 minute

Saliva Sample 2: Visit 1 on week 1. After the study exercise performed for 10 minutes.

Saliva Sample 3: Visit 2 on week 12. Before the study exercise. 0 minute

Saliva Sample 4: Visit 2 on week 12. After the study exercise performed for 10 minutes.

Enough time will be given to participants to collect the saliva.

Saliva samples will be sent to Eve Biotechnology (Calgary, Canada) for analysis of tumor suppressors and cytokines using multiplex methods.

#### Fingernails

Fingernails from the participants will be collected by self-grooming methods (participants cut their own fingernails using the clean nail clippers provided to them) at the following study visits. The fingernail will be used for proteomic analysis and cortisol measurement. At the time of study eligibility confirmation and scheduling the first visit, the participants are asked to leave the nail uncut for at least one week prior to the visits.

Fingernail Sample 1: Visit 1 on week 1. Before the study exercise.

Fingernail Sample 2: Visit 2 on week 12. Before the study exercise.

Fingernail samples will be submitted to MUSC Proteomics facility for proteomic analysis, and to Eve Biotechnology for cortisol estimation.

All the samples will be identified by the subject ID number, without any names or personal identifiers, and stored at −80oC freezer at the Radiation Oncology laboratory space (DD 329 where the PI has laboratory space). The samples will be shipped to the analytical labs on dry ice. The analysis data will be transmitted through secured web servers to the study PI and will be uploaded into REDCap for analysis by the biostatistician. Dr. Balasubramanian will oversee and coordinate the sample collection, shipment, analysis, and data collection. No genetic testing will be conducted using the samples. The samples will not be shared with any other investigators or any other purposes.

Any remaining samples will be discarded after the study is completed.

### Data Management

#### Statistical Considerations

The primary purpose of this study is to evaluate the feasibility of the group video Yogic Breathing app intervention. We will employ a parallel intervention model to compare the Yogic Breathing (YB) intervention to the control, an Attention Control (AC) intervention. After obtaining consent, participants will be randomly assigned to either group using a permuted block randomization approach; participants will be blinded to the study design as they will be assigned to one of the two mindful breathing exercises. The statistician will also be blinded to the study arms to reduce bias.

This pilot study will enroll a total of 40 study participants, 20 subjects per study arm. Given this sample size, we will be able to estimate differences between the groups within a margin of error of 14.2% with 95% confidence level. Specifically, with regards to the methods used to evaluate each outcome measure:

#### 1) Adherence (Primary Measure)

Adherence will be measured using total practice times (length and frequency). This data will be collected throughout the 12week program on all practices recorded in the app. Compliance is ensured by random virtual group visits by the study mindfulness instructor, and compliance is inherently built in with the peer support model. Adequate adherence will be defined as completion of at least 10 practices a week (10-min each), and a total of 70% of all possible sessions (126 of 180 possible sessions). We will be able to estimate this percentage within a margin of error of 14.2% with 95% confidence level.

#### 2) Acceptability (Primary Measure)

Acceptability of using the group video app to support symptom management for survivors will be evaluated using a satisfaction and system usability scale (SUS). This survey measure will be collected from all participants at study termination (Week 12). Adequate acceptability will be defined as an overall average SUS score above 68, which is well-established from a number of research studies to represent an “above average” rating of usability.

#### 3) Feasibility (Primary Measure)

Feasibility will be examined by evaluating intervention delivery factors (i.e., ease of use, barriers) and resource needs (i.e., staff time, session length, software or mobile connection challenges). Data on intervention delivery factors will be collected from participants within one month of study termination (Week 12) using a structured focus interview until saturation. Focus group interviews will be evaluated using thematic analysis with N-Vivo software (NVivo Qualitative Software, by QSR International) to identify themes and intervention implications in interview transcriptions to inform future design changes with adherence to rigorous qualitative data analysis standards. This initial theme identification process will be followed by team meetings to finalize themes and implications for the intervention design. Resource needs will be evaluated over the course of the 12-week program and serve to inform design changes needed for large-scale implementation.

#### 4) Behavioral Survey Measures (Secondary Measure)

Behavioral survey measures will be collected from all participants at study termination (Week 12) using established questionnaires and will help guide effect sizes and power calculations for the next larger-scale study (i.e., perceived stress, depression scale, anxiety scale, sleep quality measure, and MD Anderson Symptom Inventory [MDASI]). We will estimate the difference between the YB and AC groups for using a longitudinal mixed effects model. For MDASI, a 0.5 SD change is considered clinically significant. In addition, we will estimate the within-subject intra-class correlation (using an AR(1) or CS structure) and variance. These assessments will guide effect sizes and power calculations for a Phase II pivotal study.

#### 5) Focus Group Interview (Secondary Measure)

In addition to providing data on feasibility (see above), structured focus group interviews will assess the participants’ general perceptions of the intervention and technology. During these interviews, data will also be collected to identify recommendations for app modifications and improved adherence. Interview transcriptions will be evaluated using thematic analysis with N-Vivo software to identify themes and intervention implications to inform future design changes with adherence to rigorous qualitative data analysis standards. This initial theme identification process will be followed by team meetings to finalize themes and implications for the intervention design.

#### 6) Salivary and fingernails biomarkers (Exploratory Measures)

Our prior research shows salivary biomarkers alter as a result of yogic breathing. However, they were not tested in cancer patients. Therefore, to identify potential target biomarkers in cancer population that would alter as a result of yogic breathing exercise in our future large scale studies we propose performing salivary (cytokines 71plex panel, tumor suppressor DMBT1), fingernails (cortisol, and differential proteomics) analysis. Regression methods will be used to identify significant predictors among the biomarkers. Stepwise selection methods will be used for this purpose. Also, data reduction methods will be considered to find factors among the biomarkers, which may provide more parsimonious predictions in the regression model.

#### Provisions to Monitor the Data to Ensure the Safety of Subjects (if applicable)

- **Data Management and Quality Control Procedures:** The primary responsibility for data management and quality control will remain with the PI. Study project coordinators are directed to promptly report any adverse effects related to the study intervention, assessment, recruitment protocols, or any other aspects of the study to the PI, who will report these adverse effects in writing within 24 hours to both the MUSC Institutional Review Board for the Protection of Human Subjects. Following each adverse event, a thorough investigation of the circumstances leading to the event, all probable causative factors, the current status of the event, the outcome of the event, and possible course of actions to avoid such events in future will be conducted by the PIs and Co-investigators. A written summary of findings will be submitted to the MUSC IRB. The collected data will be in the form of responses to structured interviews, self-reports, and bio-behavioral assessment measures. Written forms of data will be encrypted with research numbers that are linked to participant identification number in a single Master List. This Master List is locked in the PI’s office in a secured research building. Only the PI will have keys to this file and only the project coordinator will have access to this room. The PI will review the collected data weekly to assure that all informed consent procedures are being followed, that all participant identifiers are removed from all collected questionnaires, and that all files are locked and secured. All the data entered into computer files will have only research numbers included, and no participant identifiers will be entered into digital format. The Master List of participant names and links to specific research numbers will be destroyed, and no link between identifiers and research numbers will remain upon completion of the study. Participant Safety Monitoring Procedures: All participants will be given the contact telephone number of the PI, and a 24-hour emergency contact/pager numbers. The participants will be strongly encouraged to call the PI or the emergency number if they experience any significant distress arising from study protocol. The PI will remain in contact with any participants who reported a distress. In addition, the study staff will be clearly instructed to contact the PI if any of the participants report distress due to the protocol. The PIs will then contact these participants to assess the distress. In cases of extreme distress, the PI will make a decision regarding appropriate services or referrals. All adverse effects related to the interventions and assessment procedures or any aspect of the protocol will be documented in detail and reported within the stipulated time (24 hours) to the MUSC IRB. Participant safety will be dealt with seriously at all steps of the protocol. All interview responses and data collected from the app will be uploaded into the REDCap system by the research coordinators. The MUSC Biomedical Informatics Center (BMIC) will consolidate the data, and biostatistician Dr. Ramakrishnan will analyze all data. The mobile application will be developed by PranaScience in collaboration with an information technology firm.

#### Withdrawal of Subjects (if applicable)

This is a pilot study determining the feasibility including intervention adherence. The participants will be informed that the participation is voluntary and can withdraw from the study at any time they would like by informing the study coordinator or any of the study team members. Withdrawing from this study will not affect the standard of care they receive as part of their survivorship.

#### Risks to Subjects

##### Potential Risks

YB or AC exercises do not pose any potential physical and psychological risks to participants. The training provided at beginning of study will provide participants some familiarity with exercise before they engage with program independently, to improve confidence and decrease feelings of nervousness or inadequacy. There is in-built support by the other participants (peers) within each group session. All activities whether a person does the exercise alone or in company of others is counted as practice. Dr. Hendrix and Project Coordinators (provided by SCTR, see *letter of support*) will provide additional support to participants to ensure that the subjects feel at ease and comfortable during study. The study participants will not be contacted for another study.

Some potential social risk to participants does exist, centered around threats to confidentiality and anonymity. However, by taking steps to maximizing anonymity and confidentiality we will greatly minimize such risks. These steps include removal of all personal identifiers (name, address) from collected data and replacing them with subject identification numbers. One Master List linking names with participant identification number will be secured in a locked facility and will be destroyed immediately upon completion of data collection. To improve privacy in the exercise sessions participants can rename themselves, use any makeovers (like eye masks) to modify identity, and use any Avatars to mask their identity. The App will not collect any audio or video recording of the sessions.

If the participants experience any extreme physical or mental distress secondary to participating in this study, they will be encouraged to contact the PI or a Co-Investigator immediately. At the beginning of the initial interview, the participants will be provided these contact numbers and an additional 24-hour emergency contact number. If any of the study personnel notice any adverse effects in response to, or in potential response to any project intervention, assessment protocol, or study involvement, they will immediately notify the PI. The PI will then inform the MUSC IRB within 24 hours of the initial report. It is the PI’s responsibility to address all the concerns of the participants and to set up appropriate medical referral if necessary.

##### Potential Benefits to Subjects or Others

Substantial evidence suggest that Yogic breathing contributes positively to overall health and wellbeing. Several studies indicate that both YB reduces stress and improves the quality of life in multiple patient groups including breast cancer patients and in otherwise healthy individuals. Our preliminary assessments suggest that YB leads to psychological improvements. Subjects of this study will accumulate new techniques to regulate breathing that they can practice daily at their will, leading in some cases to continuous practice that stand to provide long-term benefits. The new opportunity created through this app for social interaction with other survivors will help guide them in improved lifestyle choices and overall well-being.

##### Importance of the Knowledge to be Gained

YB is a non-invasive, non-pharmacologic, and inexpensive method. It is a time-tested intervention with documented benefits on physical and mental well-being. Additional scientific evidence is however needed to support the health claims of such holistic practices in order to reap their fullest preventive and therapeutic potential. Our study will provide initial support that a structured group video app-based YB intervention could be very useful to a) improve quality of life for survivors, and b) obtain initial data on feasibility and adherence to such an intervention. This is highly relevant for the continued wellbeing of individuals that are living with medical and psychological consequences of cancer treatment. In addition, these methods, once widely practiced, may help to prevent the onset and/or progression of future physical and mental illnesses. In summary, hundreds of thousands of Americans who have undergone cancer treatment and associated disease burden, psychological distress, mental disorders and compromised quality of life could be significantly helped by an effective, inexpensive YB adjunct therapy easily accessed via a mobile platform.

##### Sharing of Results with Subjects

The subjects who are interested in receiving copies of their data can get a copy of it at the end of the study. Their contact details and consent to contact them will be obtained at the time of informed consent process. No data belonging to other participants will be provided to any participant.

## DISCUSSION

The study results will be used to determine the feasibility, acceptability and symptom improvement characteristics of the breathing exercises when compared with the attention control intervention. In addition, pilot information on potential biomarker changes will be determined. Based on these data, a larger clinical trial will be designed to study the potential utility of the YB exercise used in the present study. Amendments to the existing protocol including change of the intervention site, inclusion criteria will be informed to the institutional review board for approval. The monetary reimbursement of $200 ($75 on visit 1 and $125 on visit 2), is used in the current study to improve retention of the trial participants. Data obtained from the current study will be used for dissemination of the information to the general public through workshops, talks and webinars by the study team. In addition, the small business entity will reach out to cancer centers and support groups and other organizations for the use of the app in cancer settings.

## Data Availability

Deidentified research data will be made publicly available when the study is completed and published.

## DECLARATIONS

### Ethics approval and consent to participate

This protocol was approved by the Institutional Review Board committees at Advarra (Pro00050387), and the Medical University of South Carolina (Pro00108511).

### Consent for publication

Not applicable

### Availability of data and materials

The data that support the findings of this study are available from PranaScience Institute but restrictions apply to the availability of these data, which were used under license for the current study, and so are not publicly available. Data are however available from the authors upon reasonable request and with permission of PranaScience Institute.

### Authors’ contributions

**S. Balasubramanian:** conceptualization, funding acquisition, methodology; principal investigator, project administration, and writing, **J.L. Harper:** study radiation oncologist, sub-award principal investigator; review & editing; **K.R. Sterba:** bio-behavioral methods, focus group interviews, conceptualization, review & editing; **R. Viswanathan:** conceptualization, study design, data curation, formal analysis, review & editing; **H. Eldredge-Hindy:** study radiation oncologist, review & editing

## Acknowledgements

SB would like to thank the support provided by Crowdlinker Inc for the app development, and the following individuals Dr. Kat Hendrix (yoga instruction), Ms. Erin Klintworth (regulatory consult), Ms. Emily Richardson (study coordination), Ms. Carley Stanley (business development), Mr. Tommy Gallien (grants and regulatory management), and Ms. Sheryl Gorsuch (fiscal management), and Dr. Graham Warren (lab support) and other members of the Department of Radiation Oncology, MUSC (administrative support).

## List of Abbreviations

AC: attention control
DMBT1: deleted in malignant brain tumor 1
ECOG: Eastern Cooperative Oncology Group
ELISA: enzyme-linked immunosorbent assay
IL: interleukin
MCP1: monocyte chemotactic protein 1
MDASI: MD Anderson Symptom Inventory
NGF: Nerve Growth Factor
SCTR: South Carolina Clinical and Translational Research Institute
SUS: system usability scale
TMP: Thirumoolar Pranayamam
YB: yogic breathing,

---

**Figure 1.**
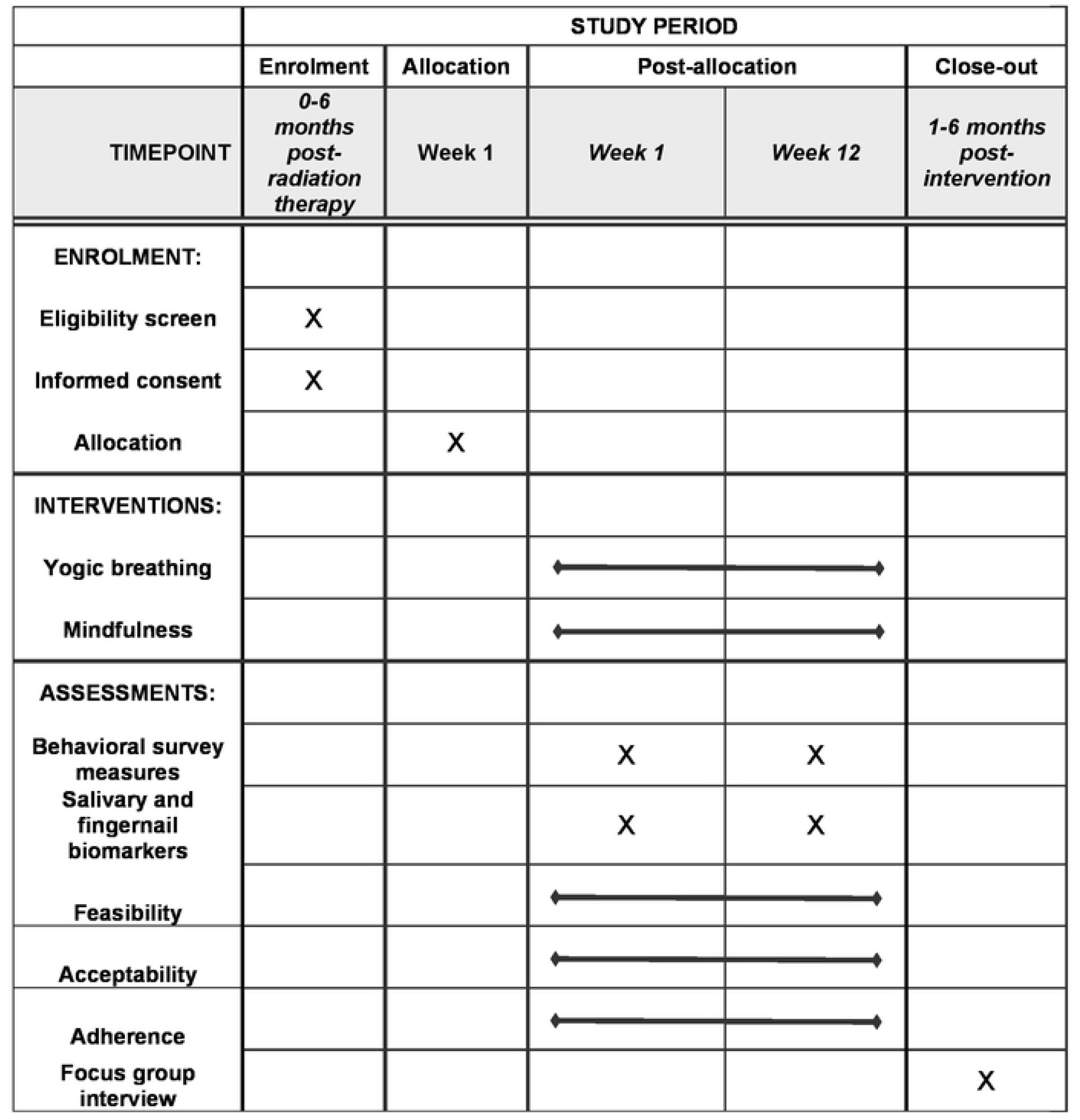
Schedule of enrolment, interventions, and assessments.

